# Comparative Impact of Individual Quarantine vs. Active Monitoring of Contacts for the Mitigation of COVID-19: a modelling study

**DOI:** 10.1101/2020.03.05.20031088

**Authors:** Corey M. Peak, Rebecca Kahn, Yonatan H. Grad, Lauren M. Childs, Ruoran Li, Marc Lipsitch, Caroline O. Buckee

## Abstract

**Background:** Voluntary individual quarantine and voluntary active monitoring of contacts are core disease control strategies for emerging infectious diseases, such as COVID-19. Given the impact of quarantine on resources and individual liberty, it is vital to assess under what conditions individual quarantine can more effectively control COVID-19 than active monitoring. As an epidemic grows, it is also important to consider when these interventions are no longer feasible, and broader mitigation measures must be implemented.

**Methods:** To estimate the comparative efficacy of these case-based interventions to control COVID-19, we fit a stochastic branching model to reported parameters for the dynamics of the disease. Specifically, we fit to the incubation period distribution and each of two sets of the serial interval distribution: a shorter one with a mean serial interval of 4.8 days and a longer one with a mean of 7.5 days. To assess variable resource settings, we consider two feasibility settings: a high feasibility setting with 90% of contacts traced, a half-day average delay in tracing and symptom recognition, and 90% effective isolation; and low feasibility setting with 50% of contacts traced, a two-day average delay, and 50% effective isolation.

**Findings:** Our results suggest that individual quarantine in high feasibility settings where at least three-quarters of infected contacts are individually quarantined contains an outbreak of COVID-19 with a short serial interval (4.8 days) 84% of the time. However, in settings where this performance is unrealistically high and the outbreak continues to grow, so too will the burden of the number of contacts traced for active monitoring or quarantine. When resources are prioritized for scalable interventions such as social distancing, we show active monitoring or individual quarantine of high-risk contacts can contribute synergistically to mitigation efforts.

**Interpretation:** Our model highlights the urgent need for more data on the serial interval and the extent of presymptomatic transmission in order to make data-driven policy decisions regarding the cost-benefit comparisons of individual quarantine vs. active monitoring of contacts. To the extent these interventions can be implemented they can help mitigate the spread of COVID-19.

**Funding:** This work was supported in part by Award Number U54GM088558 from the US National Institute Of General Medical Sciences.

## Introduction

In December 2019, Coronavirus Disease (COVID-19) emerged in Wuhan, China.^1^ It has since spread globally, reaching more than three dozen countries with over 430,000 confirmed cases by late March.^2^ To reduce further spread of the disease, governments have implemented community measures to increase social distancing for those at highest risk of infection.^3^ In China, policies include unprecedented lockdowns to reduce contacts between individuals, travel restrictions, and door-to-door temperature checks with mandatory mass quarantine.^4^

Contact tracing, a core strategy to control disease, is used to identify individuals who may have been exposed to an infectious disease and to focus interventions on this high risk group. If identified contacts are symptomatic when found, they are promptly isolated and treated in a healthcare setting. More often, contacts are found healthy, and may or may not be infected.

Depending on how much time has passed since exposure to the primary infected individual, those infected may not yet be symptomatic - this period of time between infection and symptoms is an important epidemiological trait of an infectious disease called the incubation period. How to handle these symptom-free contacts is a recurring point of confusion and controversy, particularly for emerging infectious diseases. Two essential strategies are used: voluntary individual quarantine and voluntary active monitoring. Individual quarantine involves the separation from others of an individual who is believed to be exposed to the disease, but not currently showing symptoms of it; this requires private space, provision of essentials, and investment in enforcement. A less restrictive intervention, active monitoring, involves assessing the individual for symptoms at regular intervals either through twice-daily visits by healthcare workers or phone-based self-monitoring,^5^ and, if symptoms are detected, promptly isolating the individual.

The relationship between symptoms of a disease and infectiousness to others is critical to the success of containment strategies. Previous work has found that a disease’s natural history, particularly the amount of transmission that occurs before symptom onset, greatly influences the ability to control outbreaks^6^ and the relative effectiveness of individual quarantine vs. active monitoring.^7^ Short-course diseases, such as influenza, and diseases with long periods of presymptomatic infectiousness, like hepatitis A, are impacted more strongly by quarantine than by active monitoring; however, quarantine is of limited benefit over active monitoring for the coronaviruses MERS and SARS, where persons usually show distinctive symptoms at or near the same time that they become infectious. Recent work on isolation for COVID-19 found a potentially large impact of perfect isolation, if one assumed there was limited presymptomatic transmission and a high probability of tracing contacts to be put under isolation immediately following symptom onset.^8^ A recent proposal of a contact tracing mobile phone App could allow for this instant contact tracing, decreasing the time to isolation for symptomatic contacts.^9^ Our framework enables comparison of active monitoring and individual quarantine and considers parameters such as delays, and imperfect isolation to account for known transmission of this respiratory virus after isolation in a healthcare setting.^10^

One of the key uncertainties surrounding COVID-19 is the extent of asymptomatic and presymptomatic transmission. A recent study reporting asymptomatic transmission in Germany^11^ was later found to be incorrect or misleading,^12^ adding to the confusion. There has also been uncertainty about the serial interval - the time between symptom onset of infector-infectee pairs - which in turn reflects uncertainty about the extent of presymptomatic transmission. Early estimates by Nishiura et al^13^ and Li et al^14^ were derived from limited data (24 and 6 infector-infectee pairs, respectively) and in the latter case largely reflected the prior distribution derived from SARS cases in 2003. Given the severe impact of quarantine on both resources and individual liberty, it is vital to assess under what conditions quarantine can effectively control COVID-19, and among these under what conditions it is substantially more effective than less restrictive approaches such as active monitoring, particularly given uncertainty in essential disease parameters. Here, using methods described in previous work^7^ and the latest epidemiological parameters reported for COVID-19^13,14^, we compare the ability of individual quarantine and active monitoring to reduce the effective reproductive number (*R*_*e*_) of COVID-19 to below the critical threshold of one. While mass restrictions on movements within cities have been implemented during this outbreak and are sometimes referred to as quarantines, here we focus on the effectiveness of quarantine and active monitoring on an individual basis based on contact tracing.

## Methods

### Model structure

We built upon a previously published approach^7^ using recent estimates of transmission dynamics for COVID-19 to account for critical questions and parameter uncertainties. Reflecting the uncertainty surrounding key parameters for COVID-19, we compared two sets of serial interval parameters: 1) from Nishiura et al^13^ with a mean serial interval of 4.8 days and 2) from Li et al^14^ with a mean serial interval of 7.5 days (Table 1).

**Table 1.** Disease parameters.

| Parameter | Serial interval scenario 1 |  |  | Serial interval scenario 2 |  |  |
| --- | --- | --- | --- | --- | --- | --- |
|  | Median | Mean [95% CI] | Source | Median | Mean [95% CI] | Source |
| Basic Reproductive Number ($R_0$) | 2.20 | 2.2 [1.46, 3.31] | Riou 2020 Eurosurveillanc e<sup>15</sup>; Li 2020 NEJM<sup>14</sup> | Same as scenario 1 | --- | --- |
| Serial Interval (Days) ($T_{LAT}$) | 4.6 | 4.8 [1.02, 9.81] | Nishiura 2020<sup>13</sup> | 6.99 | 7.5 [2.39, 15.48] | Li 2020 NEJM<sup>14</sup> |
| Incubation Period (Days) ($T_{INC}$) | 4.14 | 5.2 [1.11, 15.53] | Li 2020 NEJM<sup>14</sup> | Same as scenario 1 | --- | --- |
| Dispersion (k) | --- | 0.54 | Riou 2020 Eurosurveillanc e<sup>15</sup> | Same as scenario 1 | --- | --- |
| Latent period offset** (Days between latent and incubation period) ($T_{LAT} - T_{INC}$) | -0.71 | -0.77 [-1.98, 0.29] | Model Fit* | 0.59 | 0.51 [-0.77, 1.50] | Model Fit* |
| Duration of infectiousness (Days) ($d_{INF}$) | 1.8 | 2.4 [1, 6.74] | Model Fit* | 4.4 | 4.8 [1.12, 10.5] | Model Fit* |
| Relative time of peak infectiousness*** (range = 0–1) ($\beta_\tau$) | 0.38 | 0.43 [0, 0.97] | Model Fit* | 0.37 | 0.38 [0, 0.97] | Model Fit* |

Abbreviations: 95% CI = 95% Confidence Interval
\* Parameters fit via sequential Monte Carlo method
\*\* Positive values indicate symptoms before infectiousness; negative values indicate infectiousness before symptoms.
\*\*\* Value of 0 indicates linearly decreasing infectiousness; value of 0.5 indicates peak infectiousness at midpoint of duration of infectiousness; value of 1 indicates linearly increasing infectiousness.

Briefly, individuals in a stochastic branching model progress through a susceptible-exposed-infectious-recovered (SEIR) disease process focused on the early stages of epidemic growth. Upon infection, individuals progress through an incubation period (*T*_*INC*_) before onset of symptoms and a latent period (*T*_*LAT*_) before onset of infectiousness. During the duration of infectiousness (*d*_*INF*_), the relative infectiousness follows a triangular distribution (*β*_*τ*_) as a function of time *τ*since onset of infectiousness. The time offset between the latent and incubation periods (*T*_*OFFSET*_ = *T*_*LAT*_ - *T*_*INC*_), indicates presymptomatic infectiousness if negative.

During each hour of infectiousness, an individual can generate new infections (*R*_0_ * *β*_*t*_) following a negative binomial distribution with dispersion parameter (*k)*, with smaller values indicating more variability in infectiousness. Infectiousness while under individual quarantine (before symptom onset) is reduced by *γ*_*q*_ and isolation (after symptom onset) is reduced by *γ*_*i*_, a value between 0, indicating no reduction in infectiousness, or 1, indicating no transmission during that hour. Upon isolation, an individual names a defined proportion of their contacts (*P*_*CT*_), who are traced within a defined number of hours (*D*_*CT*_) and placed under either active monitoring or quarantine. Those under active monitoring are checked with a defined frequency (*D*_*SM*_), such as twice daily, and are promptly isolated if found to be symptomatic; however, prior to isolation, there is no reduction in infectiousness.

### Model parameterization

Using published values of the incubation period, the parameters for the maximum duration of infectiousness, time of peak infectiousness, and the time offset between the incubation and latent periods were fit using a sequential Monte Carlo (SMC) algorithm, also known as particle filtering.^7^ Particles with these three dimensions were resampled with an adaptive threshold to converge on a set of 2,000 that yielded simulated serial intervals that most closely match published values of the serial interval, as measured by the Kolmogorov-Smirnov (KS) test.

### Feasibility settings

As described in previous work,^7^ two settings are defined with respect to the feasibility of interventions (appendix p3). A high feasibility setting, presented as the main results, is defined as one where *P*_*CT*_ = 90% of contacts are traced and are put under either quarantine or active monitoring within half a day on average (*D*_*CT*_ = 0.5 ± 0.5 days). Contacts under active monitoring are monitored twice per day on average (*D*_*SM*_ = 0.5 ± 0.5 days). A contact under individual quarantine has infectiousness reduced by *γ*_*q*_ = 75% until symptoms emerge and prompt isolation. When symptoms emerge in a contact under quarantine or active monitoring, they are isolated in a setting that reduces infectiousness by *γ*_*i*_ = 90%, thereby greatly reducing but not eliminating infectiousness while isolated in a healthcare setting. Assuming perfect intervention performance is not possible, the high feasibility parameters represent an upper-bound on the expected ability to implement interventions based on contact tracing, and is reflected by multiple national contact investigation guidelines for COVID-19, including contact tracing within 24 hours and twice-daily monitoring for symptoms.^3,16,17^ A low feasibility setting loosens these assumptions to account for imperfect recall of who may be exposed (*P*_*CT*_ = 50%), delays in identifying or locating contacts (*D*_*CT*_ = 2 ± 2 days), infrequent or untrained monitoring of symptoms (*D*_*SM*_ = 2 ± 2 days), and imperfect isolation resulting in infections in healthcare settings (*γ*_*q*_ = 25%; *γ*_*i*_ = 50%).

### Model outputs

Unimpeded exponential epidemic growth driven by the basic reproductive number (*R*_0_) can be reduced by individual quarantine or active monitoring as measured by the effective reproductive number *R*_*IQ*_ and *R*_*AM*_, respectively. We present estimates of *R*_*IQ*_ and *R*_*AM*_ under high and low feasibility settings. The difference *R*_*AM*_ − *R*_*IQ*_ is the number of secondary cases prevented by quarantining one infected individual over active monitoring for that individual. If the prevalence of infection among traced contacts subject to quarantine or active monitoring is *p*, then the number of traced contacts who must be quarantined to prevent one secondary case relative to active monitoring is ^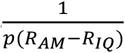^. We calculate this quantity from the model under varying assumptions about *p*, including an estimate of 0.04% estimated during SARS control in Taiwan where 24 of 55,632 quarantined contacts were found to be truly infected.^18^ To capture synergy of community-based and contact-based interventions, we measured the incremental impact of individual quarantine or active monitoring beyond community-based interventions like social distancing which we assume reduce *R*_0_.

The number of days an individual is under quarantine or active monitoring is measured as the time difference between when an individual is identified via contact tracing and when symptoms prompt isolation. We assume individuals under active monitoring or quarantine who are uninfected are followed for a duration of 14 days until clearance, consistent with recent interventions.^19^

We calculated the expected percentage of infections that result from presymptomatic infectiousness in the absence of interventions as the sum total of expected secondary cases caused by an individual prior to symptom onset divided by the total total expected secondary infections for that individual times 100. This percentage equals 0% when symptom onset precedes infectiousness and equals 100% when the entire duration of infectiousness concludes prior to symptom onset.

### Role of the funding source

The funder of the study had no role in the study design, data analysis, data interpretation, or in the writing of the manuscript. The corresponding author had full access to all of the data in the study and had final responsibility for the decision to submit for publication.

## Results

We fit the model assuming short (mean of 4.8 days; scenario 1) versus long (mean of 7.5 days; scenario 2) serial interval estimates. Model fitting by SMC for serial interval scenario 1 (mean = 4.8 days) resulted in mean duration of infectiousness 2.4 days [95% CI 1.0, 6.7]; mean time of peak relative infectiousness at 43% of the duration of infectiousness [0%, 97%]; and a mean time of infectiousness onset 0.77 days *before* symptom onset [1.98 days before, 0.29 days after] (Table 1, appendix p1). The longer serial interval in scenario 2 (mean = 7.5 days) resulted in slower disease dynamics: mean duration of infectiousness 4.8 days [95% CI 1.12, 10.5]; mean time of peak relative infectiousness at 38% of the duration of infectiousness [0%, 97%]; and a mean time of infectiousness onset 0.51 days *after* symptom onset [0.77 days before, 1.50 after] (Table 1, appendix p2). Therefore, given the same incubation period distribution with mean = 5.2 days, a serial interval with mean = 4.8 days is best fit by substantial presymptomatic infectiousness (mean = 20.5% [0%, 91.4%]) while a longer serial interval with mean = 7.5 days is best fit by limited presymptomatic infectiousness (mean = 0.065% [0%, 0.88%]).

The burden of implementing active monitoring or individual quarantine grows quickly as a function of disease incidence and the fraction of traced contacts who are not infected. Assuming uninfected contacts (who never develop symptoms) are monitored or quarantined for 14 days before clearance, the number of uninfected contacts followed grows more quickly than truly infected contacts (Figure 1). As the ratio of uninfected:infected contacts traced increases from 1:1 to 9:1, for example, the burden of uninfected contacts grows proportionally. Depending on the ratio of uninfected to infected contacts traced, individual quarantine, even if initially effective (for example, in Singapore^20^), may soon become infeasible as the epidemic grows, and will need to be supplemented or deprioritized.

**Figure 1:**
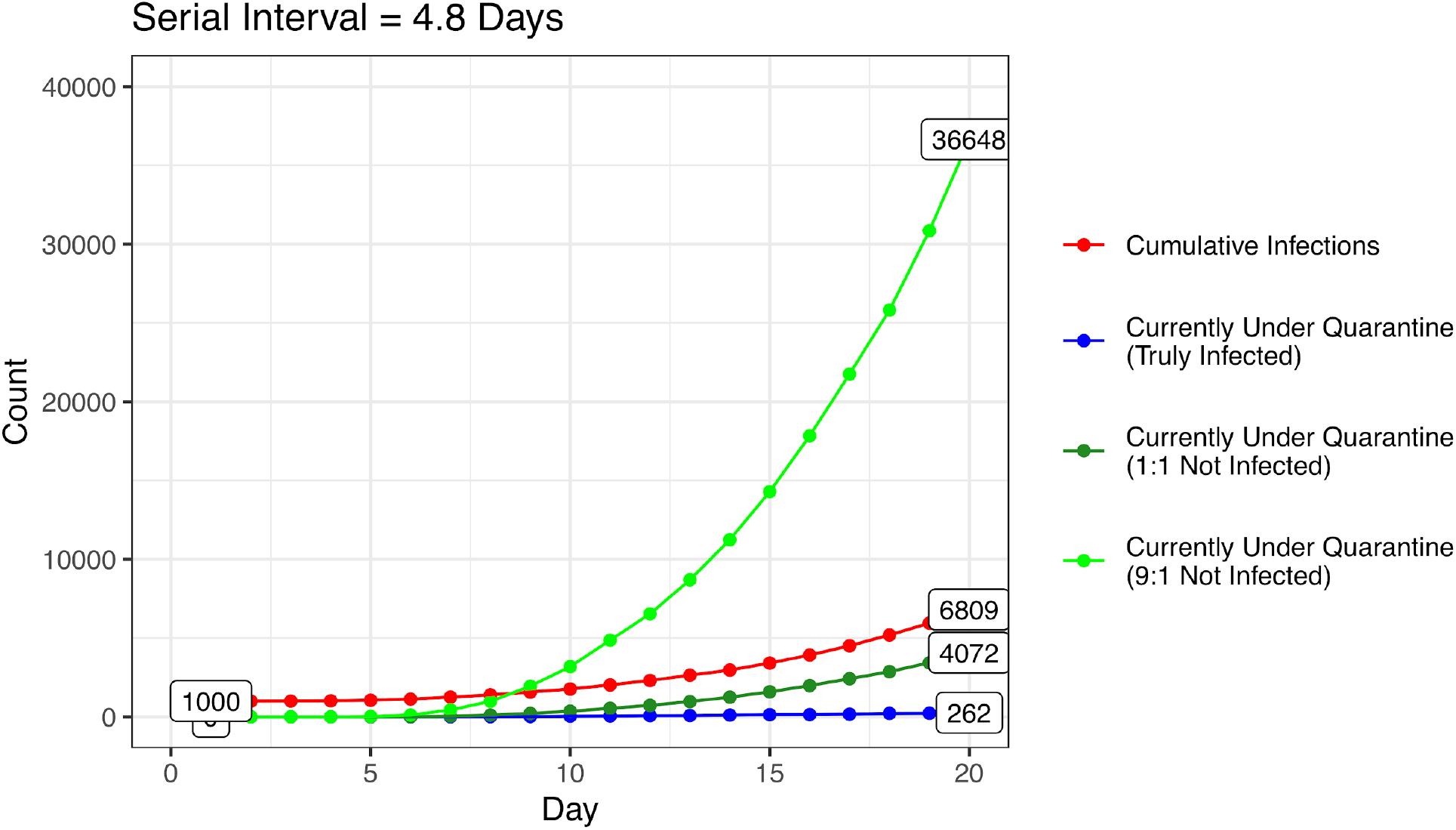
Simulated daily growth of infections and individuals under quarantine. Daily count of cumulative infections (red), truly infected contacts currently under quarantine (blue), uninfected contacts currently under quarantine assuming 1:1 ratio of uninfected to infected contacts traced (dark green, p=0.5), and uninfected contacts currently under quarantine assuming 9:1 ratio of uninfected to infected contacts traced (green, p=0.1). Model assumes interventions begin at a cumulative case count of 1,000, a low feasibility setting, *R*_0_ = 2.2, and a mean serial interval of 4.8 days.

In locations at an early stage of COVID-19, the effectiveness of individual quarantine or active monitoring depends heavily on i) aspects of the disease, especially the assumed serial interval and timing of presymptomatic transmission, and ii) aspects of the setting including the fraction of contacts traced. Briefly, a shorter serial interval, larger window of presymptomatic transmission, poor quality interventions, and a small fraction of contacts traced all reduce the ability of either intervention to decrease transmission. Unless otherwise noted, the following results focus on high feasibility settings.

First, the serial interval and extent of presymptomatic transmission are important determinants of the effectiveness of interventions. The median reproductive number was 0.57 under individual quarantine and 1.55 under active monitoring in serial interval scenario 1 (Figure 2A) and 0.49 under individual quarantine and 0.54 under active monitoring, respectively, in scenario 2 (Figure 2B). For the shorter serial interval defining scenario 1, control (*R*_*e*_ < 1) was achieved only by individual quarantine in 84% of simulations, by either intervention in 12% of simulations, and in 4% of simulations neither active monitoring nor individual quarantine reduced *R*_*e*_ < 1 in a high feasibility setting (Figure 2A). The effectiveness of active monitoring is particularly sensitive to parameter sets with earlier onset of infectiousness relative to symptoms. When *R*_0_ = 2.2, for example, *R*_*IQ*_ remains below one unless the onset of infectiousness precedes symptoms by more than two days while *R*_*AM*_ has little tolerance for presymptomatic infectiousness (Figure 3).

**Figure 2:**
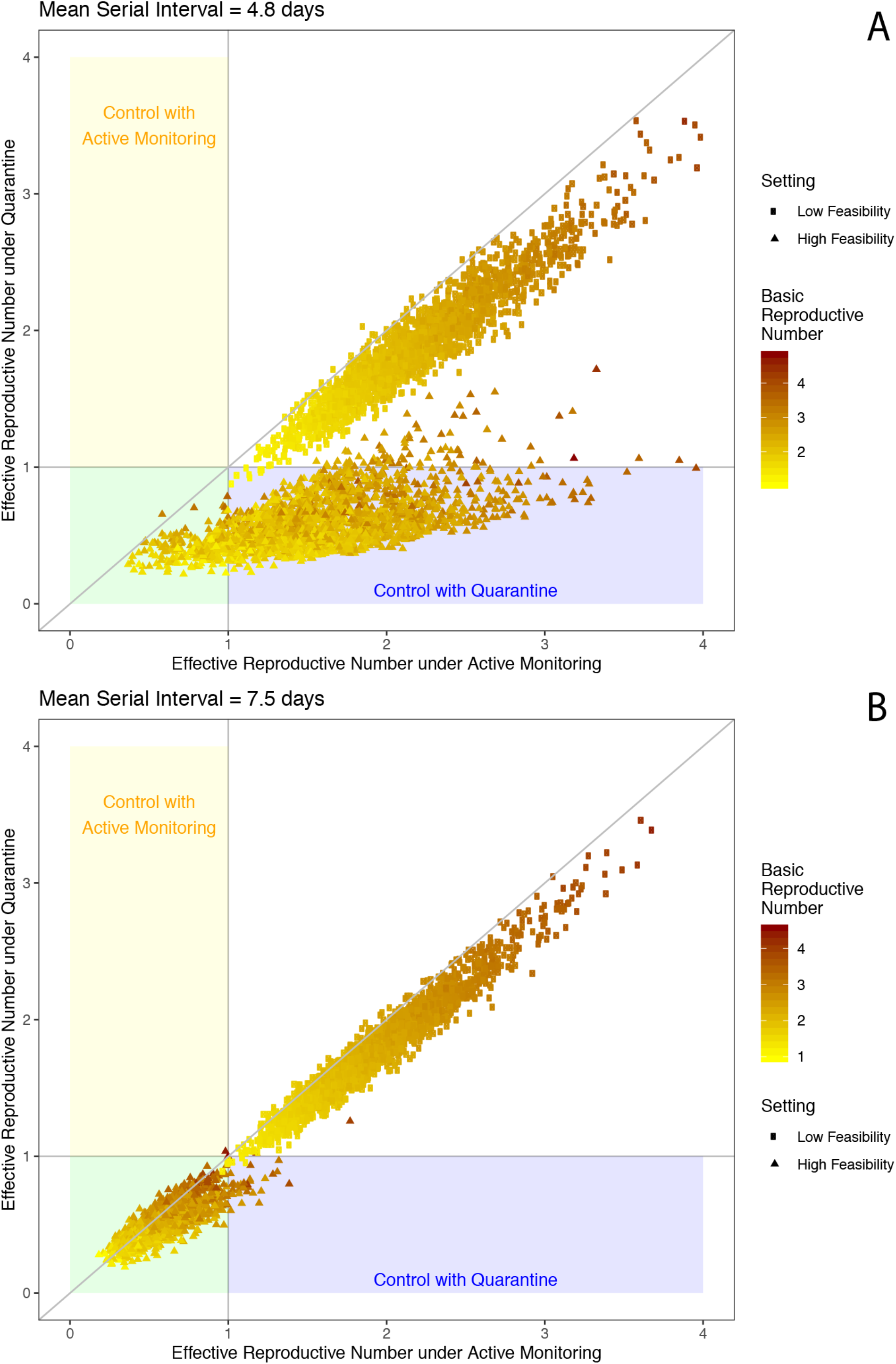
Effective reproductive number under active monitoring and individual quarantine. The effective reproductive number under active monitoring (x axis) and individual quarantine (y axis) increases with the basic reproductive number (colors) and in low feasibility settings (squares) compared to high feasibility settings (triangles) in serial interval scenario 1 (A) and scenario 2 (B). Equivalent control under individual quarantine and active monitoring would follow the y=x identify line.

**Figure 3:**
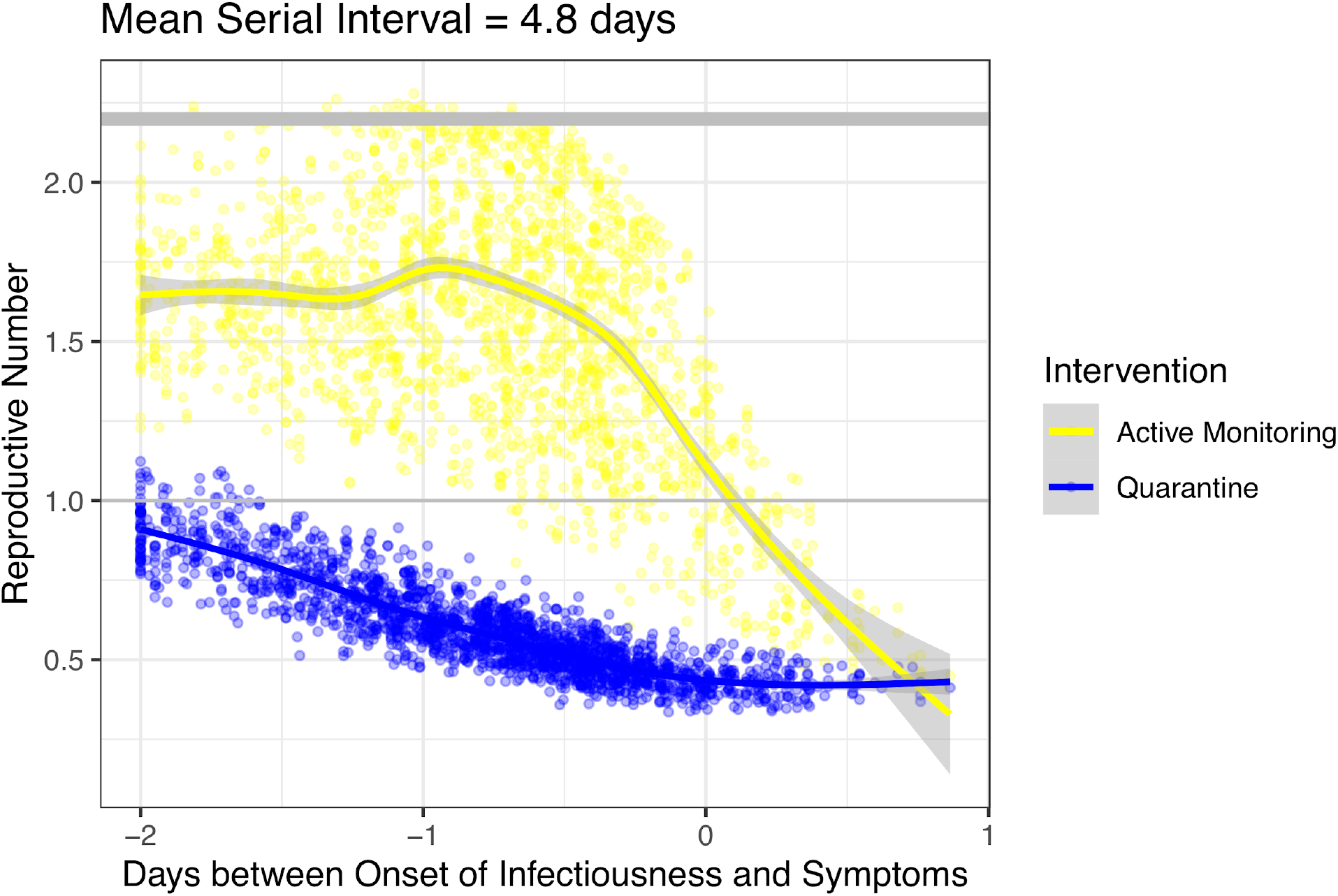
Impact of presymptomatic infectiousness on effective reproductive number. The effective reproductive number under active monitoring (yellow) and individual quarantine (blue) decreases as the extent of onset of infectiousness gets later with respect to the onset of symptoms in a high feasibility setting holding *R*_0_ constant at 2.2. An offset of −2 days indicates infectiousness precedes symptoms by 2 days, an offset of 0 days indicates onset of both simultaneously, and an offset of 1 day indicates infectiousness onset occurs 1 day after symptom onset.

In a low feasibility setting, *R*_*IQ*_ and *R*_*AM*_ remained above one, even with *R*_0_ = 1.5 (Figure 2). The fraction of contacts traced is a particularly influential consideration. As the probability of tracing an infected contact decreases, more cases are able to transmit the infection without isolation and therefore, there is a linear increase in the average *R*_*IQ*_ or *R*_*AM*_ across the population (Figure 4). Even with other operational parameters reflecting a high feasibility setting, at least three-quarters of contacts need to be traced and quarantined to reduce *R*_*e*_ < 1 in the population in the absence of other interventions. For those individuals who *are* traced and placed under active monitoring or individual quarantine, however, the impact of the interventions at reducing onward transmission by that person remains effective.

**Figure 4:**
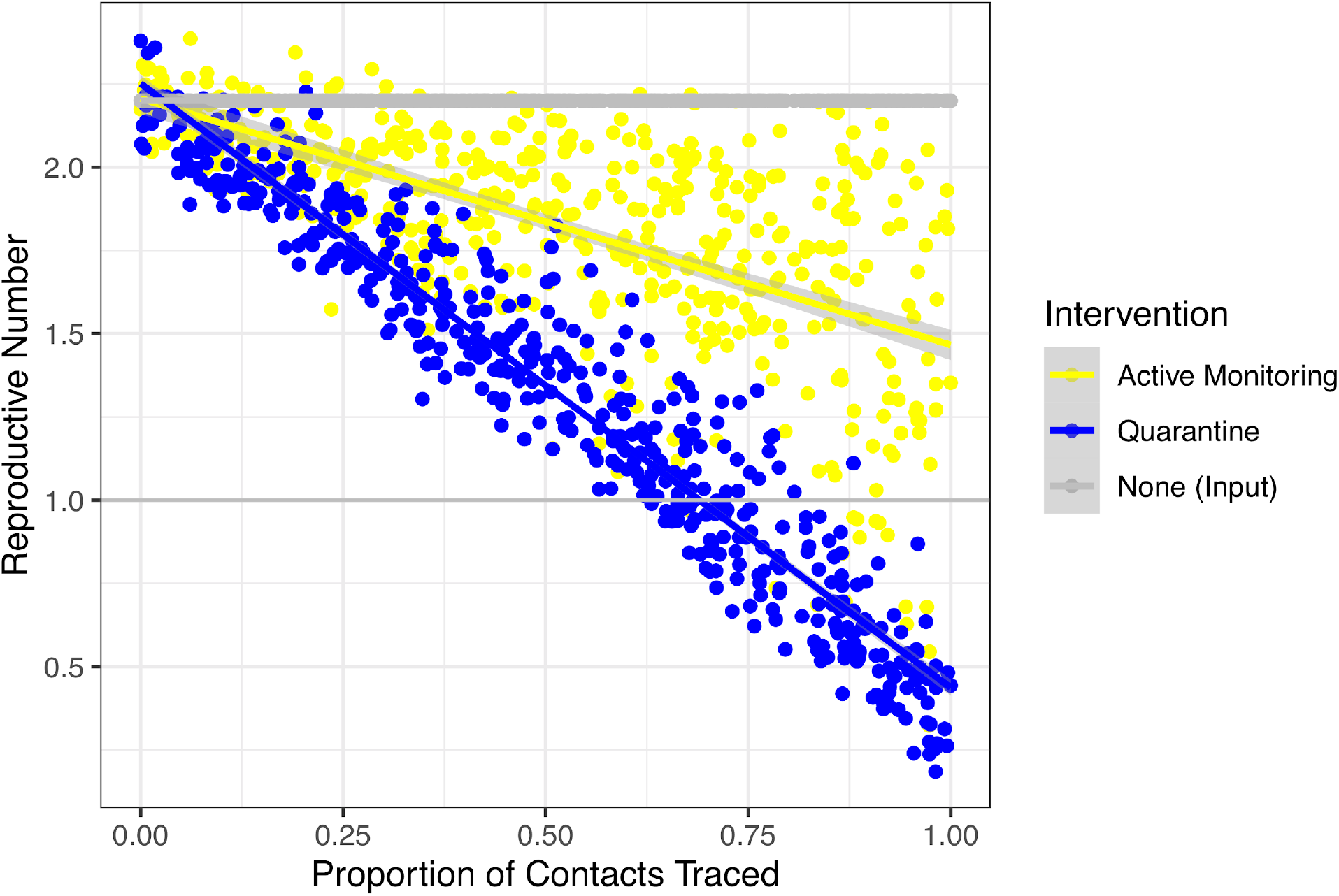
Impact of proportion of contacts traced on effective reproductive number. The effective reproductive number under active monitoring (yellow) and individual quarantine (blue) increases as the proportion of contacts traced decreases, assuming a mean serial interval of 4.8 days, and *R*_0_ = 2.2. Intervention parameters other than fraction of contacts traced are set to the high feasibility setting

In a setting where the COVID-19 case count continues to grow, resources may be prioritized for scalable community interventions such as social distancing; however, close contacts such as family members of a patient may still undergo targeted interventions. In our modeling framework, social distancing functions synergistically by reducing the reproductive number of infected individuals in the community who are not in quarantine or isolation. If social distancing reduces the reproductive number to 1.25 (e.g., 50% of person-to-person contact is removed in a setting where *R*_0_ = 2.5), active monitoring of 50% of contacts can result in overall outbreak control (ie, *R*_*e*_ < 1) (Figure 5). Tracing 10%, 50%, or 90% of contacts on top of social distancing resulted in a median reduction in *R*_*e*_ of 3.2%, 15% and 33%, respectively, for active monitoring and 5.8%, 32%, and 66%, respectively, for individual quarantine.

**Figure 5:**
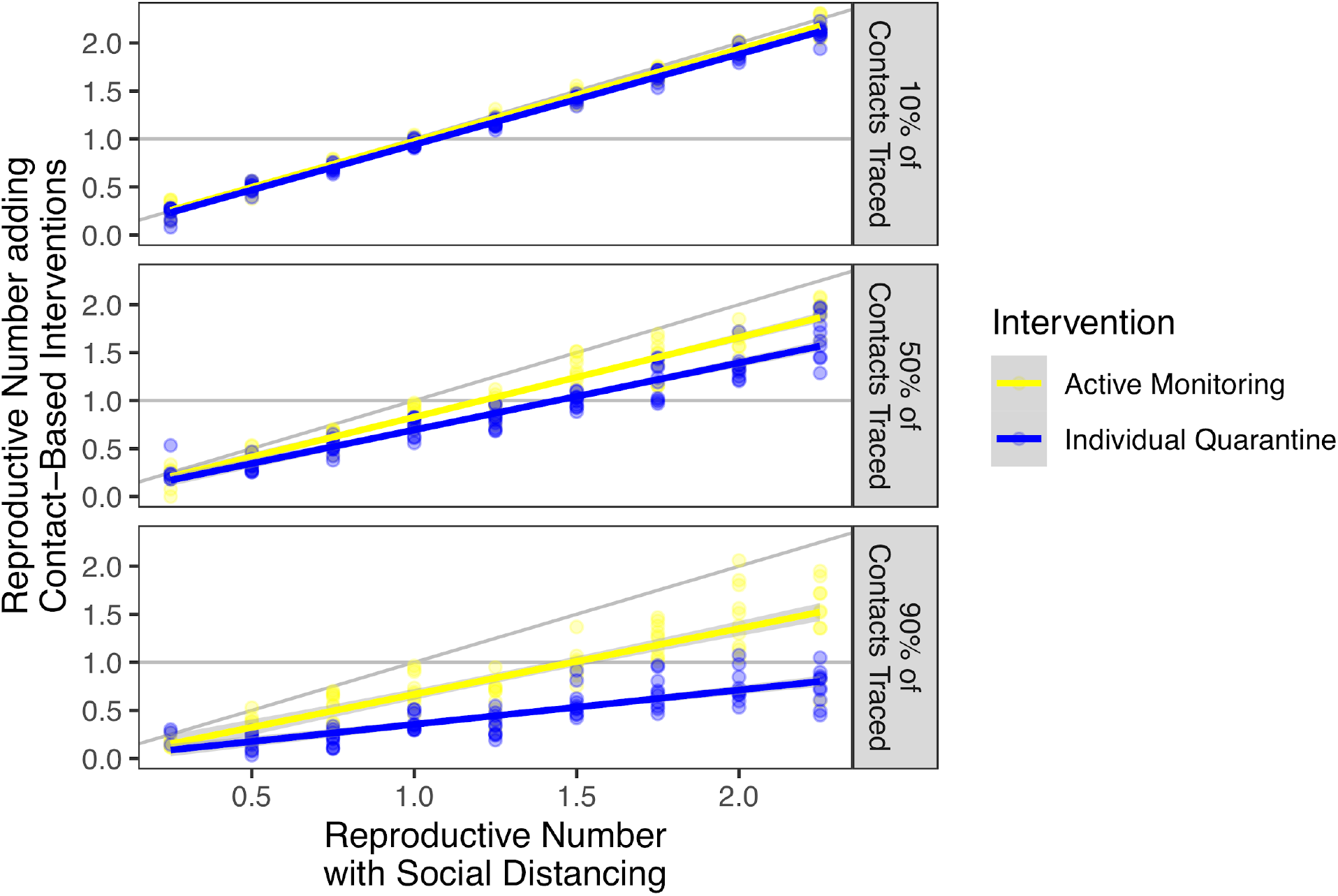
Synergistic effect of social distancing and interventions targeted by contact tracing. Active monitoring (yellow) and individual quarantine (blue) of 10%, 50%, and 90% of contacts provide an incremental benefit over social distancing for serial interval scenario 1. Intervention parameters other than the fraction of contacts traced are set to the high feasibility setting.

Given the additional cost and burden of quarantine^21^, it may be important to consider the marginal benefits of individual quarantine over active monitoring. In serial interval scenario 2 in a high feasibility setting, the median incremental benefit of individual quarantine over active monitoring was *R*_*AM*_ − *R*_*IQ*_ =0.043, which translates to a need to quarantine a median of 23 truly infected contacts to avert one infection beyond active monitoring alone. This value increases proportionally with the probability that a traced contact is not infected: 47 contacts need to be quarantined if p=50% of contacts are infected; 468 if p=5% of contacts are infected; and almost 54,000 if p=0.04% of contacts are infected as observed for SARS control in Taiwan).^18^ For the shorter serial interval scenario 1, the median incremental benefit of individual quarantine over active monitoring is much larger at 0.93, corresponding to a need to quarantine 1.1 infected contacts to prevent one secondary infection on average; though, if only p=0.04% of traced contacts are infected, nearly 2,500 individuals need to be quarantined to prevent one secondary infection relative to active monitoring.

## Discussion

To the extent that interventions based on contact tracing can be implemented, they can help mitigate the spread of COVID-19. Our results suggest that individual quarantine may contain an outbreak of COVID-19 with a short serial interval (4.8 days) only in settings with high intervention performance where at least three-quarters of infected contacts are individually quarantined. However, in settings where this performance is unrealistically high and the outbreak of COVID-19 continues to grow, so too will the burden of the number of contacts traced for active monitoring or quarantine. If the virus becomes wide-spread before any case-based control measures can be implemented and resources are prioritized for scalable interventions such as social distancing,^22^ we show active monitoring or individual quarantine of high-risk contacts can contribute synergistically with social distancing.

There may be such synergy in the data released by the WHO mission to China in February 2020.^23^ In Guangdong, WHO reports that the proportion of fever clinic testing positive for SARS-CoV-2 declined from 0.47% on 30 January to 0.02% on 16 February, during a period of intensive social distancing interventions.^23^ If one assumes that these social distancing interventions reduced transmission of nearly all infections that could cause attendance at a fever clinic, one might expect a lower total number of attendees, with proportionate declines in all infectious fever causes that are affected by social distancing. If one assumes that most fever causes are affected by social distancing, the declining proportion of SARS-CoV-2 among fever cases may reflect the benefits of interventions aimed specifically at this infection, which is to say case-based interventions like active monitoring, individual quarantine, and isolation.

The effectiveness of individual quarantine versus active monitoring, targeted by contact tracing, heavily depends on the assumptions regarding the serial interval, the amount of transmission that occurs prior to symptom onset, and the feasibility setting. Under our fitted disease natural history parameters for serial interval scenario 1, with a shorter mean serial interval and hence substantial presymptomatic infectiousness, individual quarantine was considerably more effective than active monitoring at reducing onward transmission by an infected contact. To offset this relative benefit of individual quarantine compared to active monitoring, perverse incentives to avoid quarantine would have to be correspondingly larger. Both scenarios were fit using an incubation period with mean = 5.2 days among 451 lab-confirmed cases from Wuhan;^14^ other recent estimates of the incubation period of COVID-19 include a mean of 6.4 days among 88 travelers^24^ and a median of 4 days among 291 hospitalized study patients.^25^ A shorter incubation period relative to the serial interval would be consistent with less presymptomatic transmission. On the other hand, a recent study of cases in China and Singapore found longer average incubation periods (7.1 and 9 days) and shorter serial intervals (4.6 and 4.2),^26^ which indicate an even higher proportion of presymptomatic transmission. In a scenario with these parameters, the relative benefit of individual quarantine vs. active monitoring would increase, while the total number of simulations where control is achieved under individual quarantine would decrease. The incubation period distribution, in addition to the serial interval distribution, is thus a key parameter to refine as additional information becomes available.

Under our fitted disease natural history parameters for serial interval scenario 2, with a mean serial interval of 7.5 days and hence a low amount of presymptomatic transmission, we found that both active monitoring and individual quarantine effectively reduced the expected number of secondary cases per contact below one. The incremental benefit of individual quarantine over active monitoring was minimal in this scenario, requiring hundreds or thousands of suspected contacts to be quarantined to avert one infection beyond active monitoring alone. These results suggest that with a serial interval similar to that of SARS for COVID-19, there are very few plausible conditions under which individual quarantine would offer a sufficient advantage over active monitoring to justify the substantial incremental resources required to implement individual quarantine and large incremental costs to those experiencing it. Furthermore, if the more restrictive policy of individual quarantine instead of active monitoring leads to a decrease in the percent of contacts traced, through hesitance to name contacts or avoidance of contact tracers, the small incremental benefit of individual quarantine over active monitoring in serial interval scenario 2 may cancel or invert.

Under both serial interval scenarios, in a low feasibility setting, the reproductive number was rarely brought below one under either individual quarantine or active monitoring, though a median reduction in the reproductive number of 21.0% and 13.6%, respectively for serial interval scenario 2, can still meaningfully slow the growth of an epidemic; for serial interval scenario 1, active monitoring resulted in only a 3% reduction in the reproductive number while individual quarantine resulted in a 17% reduction. If the epidemic continues to grow, however, the feasibility and social acceptability of quarantining individuals becomes a crucial consideration. In these circumstances, complementary interventions such as social distancing and pharmaceutical interventions may be needed if efficient contact tracing and rapid isolation are not readily achievable, regardless of the extent of presymptomatic transmission.

Furthermore, since contact tracing would be unable to identify contacts infected by individuals who never develop symptoms (ie, asymptomatic infectiousness rather than presymptomatic infectiousness), community interventions like social distancing are suited for mitigating transmission by asymptomatic infection while interventions targeted by contact tracing can address those exposed to individuals known to have the disease. Even if only a small proportion of infected contacts are traced, the potential transmission chains from those contacts could be prevented. The extent to which it is worth investing in imperfect contact tracing will depend on the rate of epidemic growth, which affects feasibility, and the other mix of interventions being considered.

The findings of this study are limited by the reliability of the input parameters, which are inherently uncertain during the early stages of disease emergence. The model fitting procedure is tuned to accept a wide range of inputs consistent with the published dynamics without over-fitting – thereby allowing for built-in uncertainty of input values. Additional limitations of the model include the focus on early epidemic growth in the absence of depletion of susceptibles, the assumption of consistent *R*_0_ across scenarios with different serial intervals. By assuming relative infectiousness follows a triangular distribution, we may underestimate the impact of contact-tracing interventions if relative infectiousness increases exponentially towards the end of disease instead, or overestimate the impact of relative infectious decreases exponentially after the earliest stages; however, our estimate of peak infectiousness at 38% of the duration of infectiousness suggests peak infectivity at neither end of the duration of infectiousness. By assuming the duration of infectiousness follows a uniform distribution, we may exclude long-duration shedders, which can lead to either an underestimation of the effect size, if a larger fraction of an individual’s infections happen long after isolation, or an overestimation of the effect size, if the right tail is long enough such that individuals are released from interventions and able to spark a second outbreak. In addition, our choice to consider shifts of the latent period relative to the infectious period implicitly assumes a similar shape to the underlying distributions, albeit with different means. As the amount of presymptomatic transmission will depend not only on the average timing of latent period relative to the incubation period, but also on the standard deviation of these distributions, more data on their true shapes is urgently needed.

The scope of this work compares individual quarantine and active monitoring targeted by contact tracing and does not simulate other approaches such as self-isolation or mass quarantine. In our model framework, self-isolation can be conceptualized as a scenario where all contacts are traced and under active monitoring, since recognition of symptom onset is the event that triggers isolation. Mass quarantine is expected to result in prompt isolation upon symptom onset of any truly infected individuals, but the impact of this strategy on COVID-19 will depend heavily on whether presymptomatic exposure within the group is decreased or increased by the approach to confinement. That is, mass quarantine may reduce or increase the number of uninfected contacts exposed to presymptomatic infectiousness of those who do go on to develop the disease. In serial interval scenario 1, where a mean of 20% of transmission is expected to occur before symptom onset, the positive effect of prompt isolation can be offset by an increase in presymptomatic transmission in a confined space. Mass quarantines can also result in unintended consequences that can exacerbate transmission of COVID-19 such as avoidance of contact tracers and inaccurate recall, or other infectious diseases more broadly, such as a reduction in healthcare worker support, availability of supplies, or high-density settings.^26^ The impact of travel restrictions on human mobility, a necessary first step in the causal chain to outbreak containment, is difficult to measure, but the impact has been documented in Sierra Leone during the 2014-2016 epidemic of Ebola Virus Disease.^27^

The conflicting conclusions from our two scenarios, driven largely by the differences in the extent of presymptomatic transmission, highlight the urgent need for more data to clarify key epidemiological parameters of COVID-19, particularly the serial interval and the extent of presymptomatic transmission, in order to inform response efforts. These highly influential parameters warrant further study to improve data-driven policy-making.

## Data Availability

Code will be available on github.

https://github.com/peakcm/General_Quarantine_Paper

## Contributors

All authors contributed to the study design, CMP analyzed the data, CMP and RK wrote the first draft of the manuscript, and all authors participated in the writing, reviewing, and editing of the manuscript.

## Declaration of interests

We declare no competing interests.

## Acknowledgments

This work was supported in part by Award Number U54GM088558 from the US National Institute Of General Medical Sciences. The content is solely the responsibility of the authors and does not necessarily represent the official views of the National Institute Of General Medical Sciences, the National Institutes of Health, or other contributing agencies.

## Research in context

### Evidence before this study

Two non-pharmaceutical interventions to prevent disease spread include voluntary individual quarantine and voluntary active monitoring. Previous work found that a disease’s natural history, particularly the amount of transmission that occurs before symptoms, greatly influences the ability to control outbreaks and the relative effectiveness of these two strategies. We searched PubMed Central and medRxiv for the terms “individual quarantine”, “active monitoring”, “contact tracing”, “COVID19” and “nCoV” on March 24, 2020, with no search date or language restrictions. We identified several studies reporting estimates of epidemiologic parameters of COVID-19, as well as others focused on measures to control the COVID-19 outbreak. However, few focused specifically on contact-based measures. Recent work on isolation for the 2019 Coronavirus Disease (COVID-19) found a potentially large impact of perfect isolation, if one assumed there was limited pre-symptomatic transmission and a high probability of tracing contacts to be put under isolation immediately following symptom onset. However, the estimates for the serial interval of COVID-19, which impacts the amount of pre-symptomatic transmission, are varied.

### Added value of this study

As COVID-19 continues to spread, better understanding how to contain it becomes critical. Here, using methods we previously developed and the latest epidemiological parameters reported for COVID-19, we compare the ability of individual quarantine and active monitoring to reduce the effective reproductive number of COVID-19 to below the critical threshold of one. We provide an estimate of presymptomatic transmission specifically for COVID-19, a key parameter for understanding outbreak dynamics. We further develop a metric for the feasibility of scaling of active monitoring and individual quarantine and examine the synergistic effect of contact-tracing interventions with social distancing, which can guide public health response to this pandemic.

### Implications of all the available evidence

Assuming a reported serial interval of mean 4.8 days, the incremental benefit of individual quarantine over active monitoring was substantial as a result of faster dynamics and more presymptomatic transmission. However, using a SARS-like serial interval of mean 7.5 days, individual quarantine and active monitoring are similarly effective at controlling onward transmission in a high feasibility setting. The burden of placing uninfected contacts under individual quarantine can grow untenable due to a longer duration in quarantine before clearance (assumed 14 days) and a high ratio of uninfected contacts traced per truly infected contact. In such settings, resources may be prioritized for broader social distancing measures, and active monitoring or individual quarantine of high-risk contacts can contribute synergistically. The sensitivity of these results to the estimated serial interval highlights the urgent need for better data to guide policy decisions.

## Appendix

**Figure S1:**
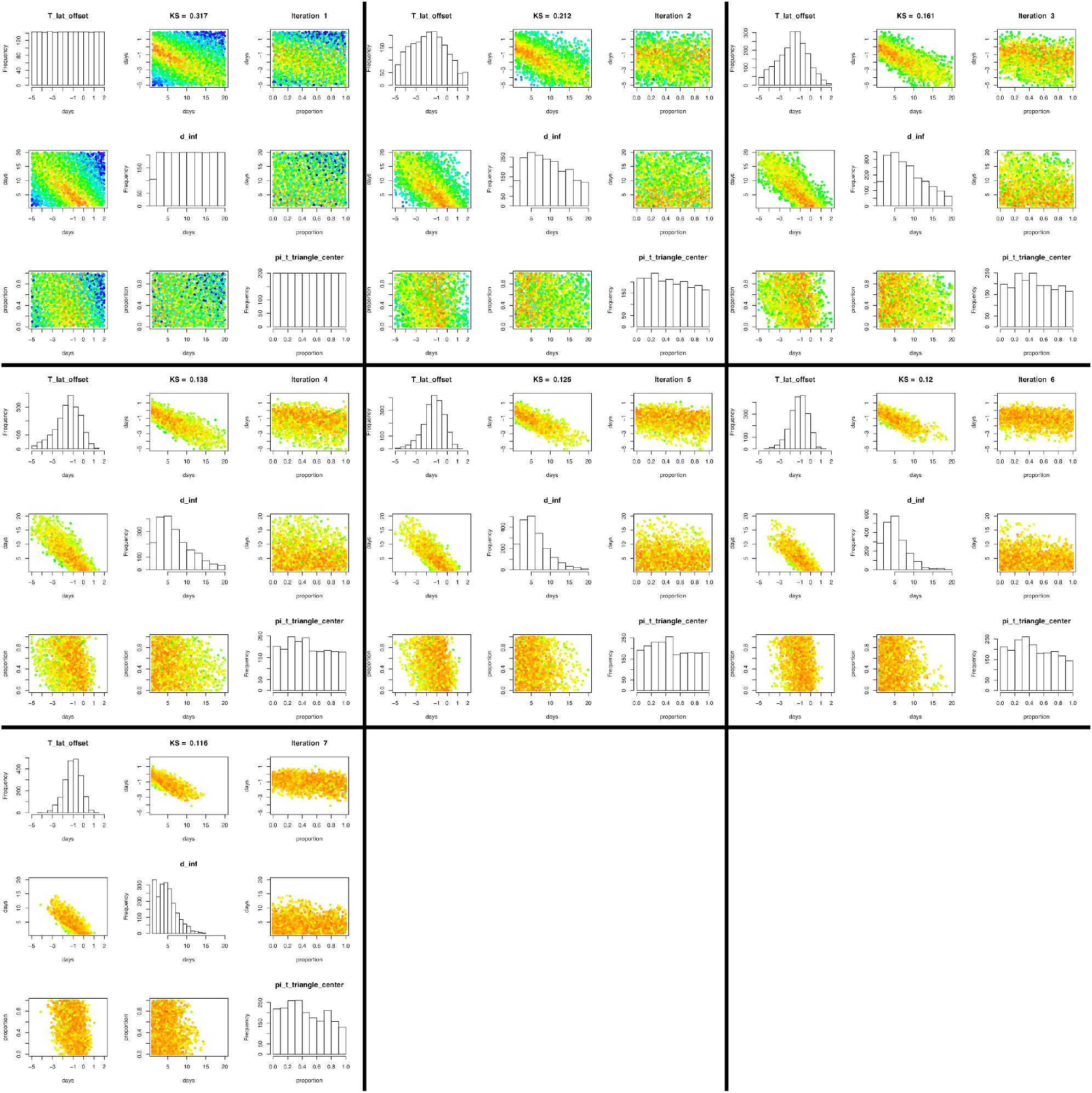
Parameters fit to serial interval scenario 1. Univariate histograms and bivariate heatmaps for each of three input parameters in serial interval scenario 1: the time offset between the latent and incubation periods (*T*_*OFFSET*_); maximum duration of infectiousness (*d*_*INF*_); and time of relative peak infectiousness (*β*_*τ*_). Convergence by sequential monte carlo (SMC) in iteration 7 with median Kolmogorov-Smirnov test statistic KS = 0.116.

**Figure S2:**
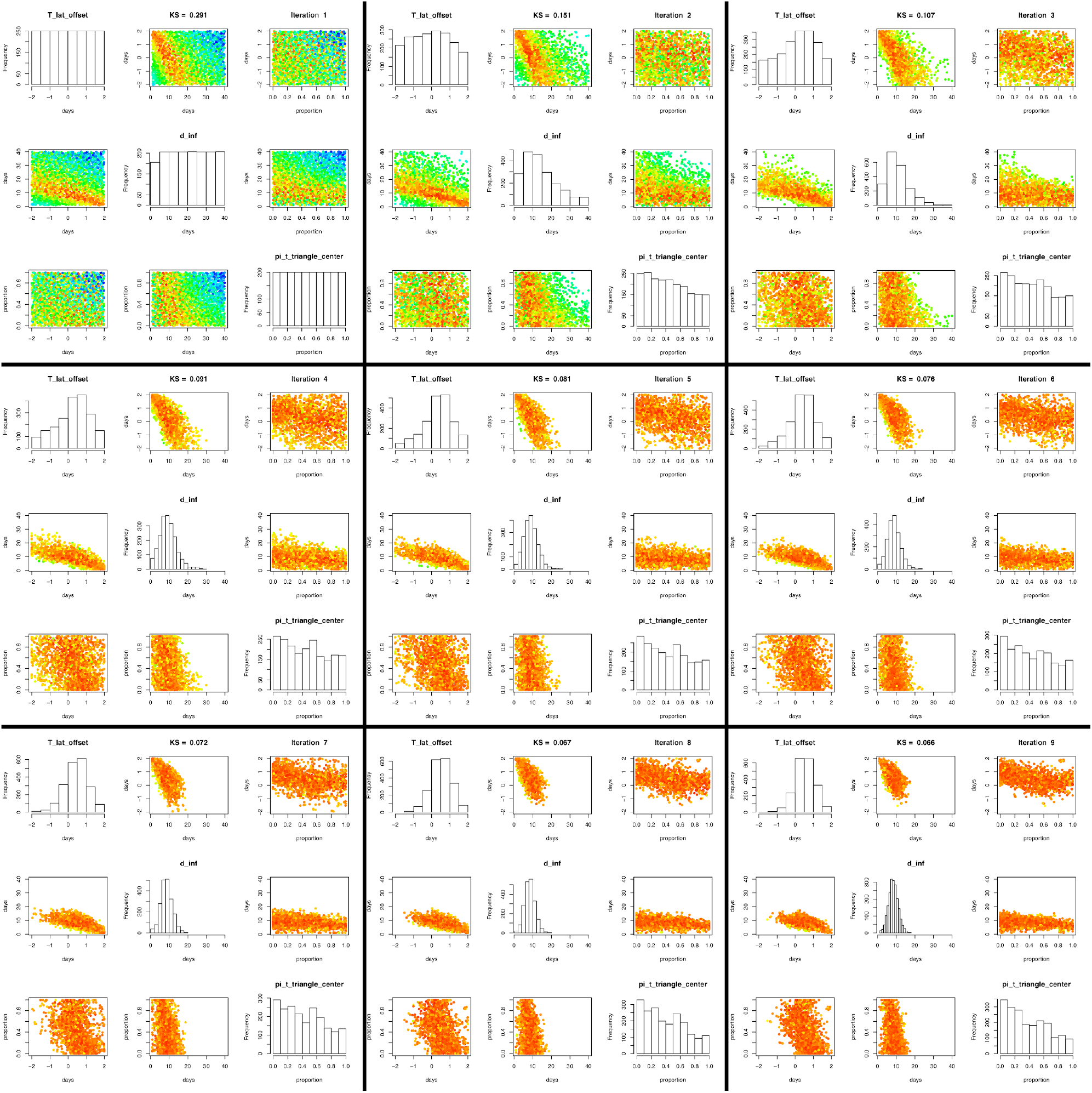
Parameters fit to serial interval scenario 2. Univariate histograms and bivariate heatmaps for each of three input parameters in serial interval scenario 2: the time offset between the latent and incubation periods (*T*_*OFFSET*_); maximum duration of infectiousness (*D*_*INF*_); and time of relative peak infectiousness (*β*_*τ*_). Convergence by sequential monte carlo (SMC) in iteration 7 with median Kolmogorov-Smirnov test statistic KS = 0.066.

**Table S1.** Intervention Parameters.

| Parameter | High feasibility setting | Low feasibility setting |
| --- | --- | --- |
| Probability of tracing an infected contact ($P_{CT}$) | 0.9 | 0.5 |
| Delay in tracing a contact ($D_{CT}$) | $0.5 \pm 0.5$ days | $2 \pm 2$ days |
| Reduction in infectiousness during quarantine (*for pre-symptomatic contacts under quarantine*) ($\gamma_q$) | 0.75 | 0.25 |
| Frequency of monitoring symptoms (*for pre-symptomatic contacts under active monitoring*) ($D_{SM}$) | $0.5 \pm 0.5$ days | $2 \pm 2$ days |
| Reduction in infectiousness during isolation ($\gamma_i$) | 0.9 | 0.5 |

## Notes

### Competing Interest Statement

ML reports grants from NIH/NIGMS, during the conduct of the study; personal fees from Merck, grants from CDC, grants from Open Philanthropy Project, outside the submitted work.

